# Investigating the Psychophysiological Effects of a Telehealth-enabled Multi-sensory Environment on Anxiety among Young Adults

**DOI:** 10.64898/2026.03.08.26347897

**Authors:** Roxana Jafarifiroozabadi, Namgyun Kim, Hani Patel, Jaehoon Lee, Stephen N. Parker

## Abstract

Anxiety symptoms among adults in the U.S. have increased significantly in recent years, with higher prevalence among younger adults. This industry–academia collaborative study evaluates the effectiveness of a telehealth-enabled multi-sensory environment as an integrated intervention on reducing anxiety levels among young adults. A sample of 30 participants was recruited from a university population in the U.S. Anxiety levels were assessed during three five-minute episodes: baseline, exposure to Trier Social Stress Test (TSST), and physical sensory environment with telehealth (pre-recorded guided meditation) as the intervention. Physiological data—electrodermal activity (EDA), number and duration of eye fixations and saccades— were collected continuously using sensors (EmbracePlus) and eye-tracking (Tobii Pro Glasses). Subjective data were measured using the State-Trait Anxiety Inventory (STAI) and semi-structured exit interviews. Pairwise comparisons based on complete datasets from 25 participants revealed a significant decrease in EDA (*P* <.001), mean frequency of saccades (*P* =.011) and eye fixations (*P* <.001), and perceived state anxiety scores (*P* <.001) among participants following TSST, indicating the effectiveness of the telehealth-enabled multi-sensory environment in anxiety reduction. Semi-structured interviews also highlighted participants’ preferences regarding key sensory environment features, including tactile, form, lighting, furniture types, and configurations. Findings from this study will inform the design and implementation of telehealth-enabled multi-sensory environments in the future educational settings to improve anxiety symptoms among young adults.

## 1. Introduction

Young adults are experiencing rising rates of mental health problems, including anxiety, during their academic journey in university environments (Arun et al., 2017; Okechukwu et al., 2022). To address these challenges, universities often provide mental health support with various counselling services, such as telehealth via online platforms, peer-support programs, or wellness initiatives (Grégoire et al., 2024). While these services can be effective in reducing anxiety, they commonly remain insufficient to address the various needs of young adult students due to long wait times, limited access to resources, limited staff availability, and stigma around seeking help from the mental health services on the campus (Brown et al., 2023; Daniel Eisenberg et al., 2021; Dunley & Papadopoulos, 2019; Ebert et al., 2019). Thus, **there is a need for alternative mental and behavioral health interventions on campus that are accessible, scalable, and non-stigmatizing while supporting anxiety reduction and emotional regulation among young adults.**

In addition, the existing literature demonstrates that multi-sensory environments can positively impact measurable physiological or psychological outcomes, such as anxiety, among individuals across healthcare, education, and public settings (Collier & Jakob, 2017; Dorn et al., 2020; Haig & Wagstaf, 2024; Watchorn et al., 2025; West et al., 2017). Emerging evidence highlights that multi-sensory environments incorporating elements, such as lighting, sound, texture, furniture, scent, and tactile materials that engage multi-sensory systems in human body can lead to a significant reduction in agitation and distress and increase calmness, feeling of safety, and development of coping strategies among individuals in healthcare environments (Dawson et al., 2025; Forsyth & Trevarrow, 2018; Haig & Wagstaf, 2024; Kjær et al., 2024) (McTeague et al., 2025; Xu et al., 2025). In educational settings, one study examined a multi-sensory room environment with interactive features among 75 nursing students. Data collected through surveys showed participants’ satisfaction with emotional wellbeing and academic readiness following experiencing the environment (Bruce et al., 2024). Another study used randomized controlled trials and reported that the multi-sensory environments enhanced vagal (parasympathetic) regulation among thirty-nine healthy young adults (university students) following a thirty-minute exposure to the environment, suggesting broad applicability for mental health support in higher-education settings (Otsuka et al., 2025).While these studies emphasize the overall implementation efficacy of multi-sensory environments in educational settings, including university campuses, **little evidence exists on the impact of integrating technological interventions (e.g., telehealth) within these environments or varying exposure durations on anxiety levels on young adults.**

Moreover, studies have examined various objective and subjective indicators of anxiety when assessing the impact of the environment on individuals’ anxiety levels. Objective measures of anxiety, including eye movement metrics (e.g., saccades and fixations) and electrodermal activity (EDA), have been widely adopted as objective measures of individuals’ emotional and physiological arousal in the existing literature (Braithwaite et al., 2013; Clauss et al., 2022; Winz et al., 2022). Prior studies have reported that individuals in heightened anxiety states tend to exhibit more frequent and rapid saccadic movements (Agustianto et al., 2025; Manoli et al., 2021) and shorter eye fixation durations, whereas lower anxiety or more relaxed conditions are associated with longer fixation durations, indicating more stable and sustained visual attention (Konovalova et al., 2021). Additionally, previous studies have utilized EDA (changes in the electrical conductance of the skin) to detect changes in individuals’ mental and emotional states across diverse contexts, including stress, cognitive workload, and affective responses (Kim et al., 2020). EDA has shown stronger performance than measures such as heart rate or skin temperature in distinguishing baseline from social anxiety states (Shaukat-Jali et al., 2021). Subjective measures of anxiety are typically collected through self-reported questionnaires, such as the Spielberg’s State-Trait Anxiety Inventory (STAI) which is considered “gold standard” for measuring two distinct anxiety concepts: state anxiety (STAI-S), used to indicate how they feel at a particular moment in time, and trait anxiety (STAI-T), reflecting how individuals generally feel (Spielberger, 2012). The tool is also used to establish a ground truth for anxiety levels when investigating the physiological indicators of anxiety (Nath & Thapliyal, 2021; Tadayon et al., 2018); however, **it remains unclear which measures are most effective for evaluating anxiety among young adults and the extent to which subjective and objective indicators of anxiety align during such assessments.**

To address the research gaps mentioned above, this study extends the current knowledge by investigating the effectiveness of a multi-sensory environment with telehealth as an integrated intervention on anxiety levels among young adults on a university campus. The study will explore: Q1. What is the effect of a telehealth-enabled multi-sensory environment on state anxiety levels from pre-to post-exposure among young adults? Q2. To what extent does a brief exposure (approximately five minutes) to a telehealth-enabled multi-sensory room influence state anxiety levels among young adults? Q3. To what extent a combination of objective and subjective indicators of anxiety (eye movements, EDA, STAI-S) can determine anxiety levels among young adults?

## 2. Methods

The research questions were investigated using a single-group quasi-experimental study using a within-subjects (repeated-measures) design in which participants served as their own controls. The overall methodology is described in the following sections.

### 2.1 Design and development of the multi-sensory environment prototype

The original concept for the prototype was adopted from a pop-up prototype for sensory-enabled architecture designed to promote mental health, relaxation, and wellness (Parker & Morgan, 2025). In collaboration with the Texas A&M Arch4Health Team, the prototype was designed to be implemented in the lobby space of the Department of Architecture building on a university campus located in Texas, U.S., to provide accessible telehealth interventions for young adult educators in the Department (Figure 1). Using neuro-inclusive design and trauma-informed design as guiding design frameworks, the prototype integrated tactile, visual, and kinetic sensory elements to engage multiple senses—sight and touch—to create a holistic experience and a sense of comfort and safety while allowing them to control and personalize their environment. Figure 1 indicates the development and assembly of the prototype. Additional information regarding the specific design features implemented in the prototype is demonstrated in Table 1.

**Figure 1.**
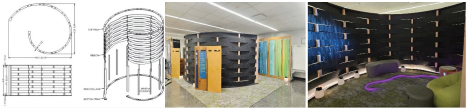
The conceptual design, installation, and assembly of the multi-sensory environment in the lobby space on of the university campus

**Table 1.** Design features implemented in the prototype.

| Environmental Feature | Description | Trauma-Informed Design Principle(s) |
| --- | --- | --- |
| **Custom enclosures (cloisters/pods)** | Provide privacy while allowing passive observation; acoustically and visually calming environment | Safety, Security, and Privacy; Hope, Dignity, and Self-esteem |
| **Dynamic furniture (e.g., bean bag chairs)** | Designed for behavioral health; allows for movement, safety, and comfort | Safety, Security, and Privacy |
| **Lighting Features** | Static lighting quality with the following fixtures:<br>1. Flexible LED strip lighting – New Linaire Mini 3D spec attached; 3000K<br>2. RGB LED flexible strip (LED Twinkle Fiber Optic Lights Star Ceiling Kit): 10 Watts<br>3. Rechargeable LED lanterns with opal diffuser; 2700K; ~100 lumens each<br>4. Overhead Ambient (Ceiling Grid Lighting); Recessed troffer or panel LED; 3500K | Empowerment and Personal Control |
| **Tactile stimulation opportunities** | Textured materials for touch-based sensory regulation, including the acoustic Polyethylene Terephthalate (PET) felt panels for the curved walls. | Empowerment, Safety Joy, Beauty, and Meaning |

### 2.2 Sample

An overall sample of 30 young adults (ages 18–25) were recruited from a university population located in the U.S., through purposive sampling. Power analysis was conducted using RStudio (Version 2024.12.1) to determine the required sample size for detecting a medium effect size (f = 0.25) with a significance level of α = 0.05 and a desired power of 0.80. Participants were invited to join the study via emails. Consent forms were obtained prior to initiating the study, and participants were asked to complete a screening questionnaire to collect data on their demographics, medical and mental health history, prior experience with telehealth interventions prior to their participation in the study. Exclusion criteria included: 1) being actively under the care of a psychiatrist for anxiety or having a history of admission to a psychiatric facility; and 2) inability to speak English.

### 2.3 Study Procedure

Following the participant recruitment and screening process, objective and subjective anxiety indicators (Table 2) were captured during three five-minute episodes (Tarrant et al., 2018): Episode 1: Baseline data were collected with participants resting in a seat with eyes open upon arrival, Episode 2: Participants underwent a standardized stress-inducing task using the Trier Social Stress Test (TSST) (Birkett, 2011) which requires them to make an interview-style presentation without receiving any feedback or encouragement from the research team (Kirschbaum et al., 1993). Episode 3: Participants were assigned to receive a five-minute, pre-recorded telehealth care session via noise-cancelling headphones in the multi-sensory environment prototype (intervention). The telehealth intervention was a five-minute mindfulness exercise created by the University of California Los Angeles (UCLA) Mindfulness Education Center, advancing evidence-based mindfulness programs to promote stress reduction, resilience, and overall well-being for individuals including young adults on university campuses (Winston, n.d.). Data related to objective indicators of anxiety, including electrodermal activity (EDA), frequency and duration of eye fixations and frequency of saccades were collected continuously during the episodes. EDA data were collected continuously using wrist-mounted, wearable EDA sensors at microsecond-level resolution and were sampled at 4 Hz. Continuous eye movement data were collected using the Tobii Pro Glasses 3, a wireless eye-tracking system, with a sampling rate of 100 Hz and obtained through the Tobii Pro Lab (Version 25.7). The online version of the State Anxiety Inventory of the State-Trait Anxiety Inventory (STAI) was administered to participants via iPad Pro at the end of each five-minute episodes to collect self-report scales measuring state anxiety, requiring participants to indicate how they feel at a particular moment in time. The Semi-structured exit interviews were also conducted following participants’ exposure to the telehealth intervention in the multi-sensory environment prototype to obtain participants’ feedback on their experience.

**Table 2.** Objective and subjective anxiety indicators.

| Outcome of Interest | Variable Type | Measures | Method | Analytical Strategy |
| --- | --- | --- | --- | --- |
| Anxiety Levels | Objective | *Electrodermal Activity (EDA)*: Changes in the electrical resistance of the skin. EDA has a tonic and phasic components. The tonic component accounts for slow-changing components and the background characteristics of the signal (skin conductance level: SCL) while phasic components are the faster-changing elements of the signal (skin conductance response: SCR) (Posada-Quintero et al., 2018; Sarchiapone et al., 2018). | EmbracePlus (Empatica) wristwatch | One-way repeated-measures analysis of variance, Pairwise comparisons via RStudio (Version 2024.12.1) |
|  |  | *Eye-tracking*:<br>*Fixation duration*: How long participants fixate on the sensory features available within the environment (anxiety is linked with shorter fixation durations on stimuli in avoidance patterns, or longer fixations in hypervigilance patterns)<br>*Frequency of fixations*: Scanning vs. avoidance tendencies.<br>*Frequency of Saccades*: Rapid, ballistic eye movements that abruptly change the point of fixation. | Eye-tracking sensor/glasses: Tobii Pro Glasses 3 |  |
|  | Subjective | *State-Trait Anxiety Inventory* (Metzger, 1976): A validated tool with 20 items for assessing trait anxiety (how participants generally feel) and 20 for state anxiety (STAI-S), indicating how participants feel at a particular moment. The STAI-S questionnaire contains question fields such as “I feel calm”, “I feel upset”, etc. which can be rated on a 4-point scale (not at all, somewhat, moderately so, very much so). The total score ranges from 20 to 80, with scores of 20–37 indicating low or no anxiety, 38–44 indicating moderate anxiety, and 45–80 indicating high anxiety (Spielberger, 2012). | Online survey (administered via an iPad Pro) |  |
|  |  | Semi-structured Interviews: Questioned explored overall experience and participant preferences regarding the study intervention. | Audio-recorded and verbatim-transcribed interviews | Content analysis via MAXQDA (2024) |

### 2.4 Analytical Strategy

Prior to analysis, data processing was conducted through the following methods:

#### 2.4.1 EDA (Tonic)

Continuous EDA data, representing the tonic activity, were measured in microsiemens (µS) and sampled at microsecond resolution. Noises and artifacts in the EDA data were removed using a Blackman window filter with twelve data points per block while maintaining signal integrity (Podder et al., 2014; Posada-Quintero et al., 2018). Following the smoothing process, the EDA signals were normalized at the individual level to account for inter-individual variability in baseline skin conductance levels. The normalized signals were then segmented according to the timestamps of each experimental episode to enable comparative analysis of changes in the tonic component across study conditions. Data quality checks and visual inspections were subsequently performed to identify extreme or non-responsive patterns that could disproportionately influence the results. Datasets exhibiting unusually large normalized tonic values or atypical response patterns across sessions were considered outliers and excluded from the final analysis to ensure the robustness and reliability of the statistical comparisons.

#### 2.4.2 Eye movement (Saccadic movements and Fixations)

Raw eye movement data were recorded via sampling frequency of 100 Hz and obtained from the Tobii Pro Lab (Onkhar et al., 2023). Periods of missing gaze data caused by blinks were automatically detected. Short signal losses were interpolated to preserve temporal continuity, whereas longer gaps were classified as blinks and excluded from further analysis. Fixations and saccadic movements were identified using a velocity-threshold identification (I-VT) algorithm. Gaze samples exceeding a predefined angular velocity threshold (30°/s) were classified as saccades, whereas consecutive samples below the threshold were grouped as fixations if sustained for a minimum duration (200ms). To examine differences across episodes, this study used the total number of saccadic movements and fixations detected during each episode. The average fixation duration within each episode was calculated and used as an additional metric to assess changes in visual attention patterns across conditions.

Following data processing, one-way repeated measures ANOVA and post-hoc pairwise comparisons were used to evaluate pre–post exposure and between-episode differences for physiological indicators of anxiety. Nonparametric data obtained from the State Anxiety Inventory (20 items) were analyzed and compared across the episodes using the Friedman tests with post-hoc Wilcoxon signed-rank tests (Bonferroni-corrected). Audio-recorded semi-structured interviews were transcribed verbatim. Content analysis was conducted via MAXQDA (2022). Descriptive statistics, including means, standard deviations, and percentages were used to describe the data. All analyses were conducted using via RStudio (Version 2024.12.1).

## 3. Results

### 3.1 Participants

A total of thirty participants completed this study. The sample consisted of 60% female and 40% male, comprising twenty-one participants aged between (18-22), and nine participants aged were 23 and above. Five participants had the history of mental illness in one or more areas, includes (Depression, Anxiety, and ADHD) out of which one also had a history of the suicide ideation/thoughts and two participants were receiving care from the therapist or clinician for the anxiety. None of the participants had a history of hospitalization in a psychiatric facility. The majority of participants’ height and weight (N = 23) ranged between (5’-2” to 5’9”) and (120-199 lbs). None of the participants were pregnant at the time of the study. Also, 16 participants wore corrective lenses and 23 mentioned English as their first language. Considering the inclusion and exclusion criteria, missing data points, and outliers, analysis was conducted for only twenty-five participants in this study (Table 3).

**Table 3.** Demographics for participants who completed the study.

| Participants Characteristics | Values | (Counts %) |
| --- | --- | --- |
| **Total participant =30** |  |  |
| **Gender** |  |  |
| Male | 12 | 40.00% |
| Female | 18 | 60.00% |
| **Age Range** |  |  |
| 18-22 | 21 | 70.00% |
| 23-25 | 7 | 23.33% |
| 26 and above | 2 | 6.67% |
| **Height** |  |  |
| Under 5' 2" | 1 | 3.33% |
| 5'-2" to 5'-5" | 10 | 33.33% |
| 5'-6" to 5'-9" | 13 | 43.33% |
| 5'-10" to 6'-1" | 5 | 16.67% |
| 6'-2" or taller | 1 | 3.33% |
| **Weight** |  |  |
| Under 120 lbs | 6 | 20.00% |
| 120 – 159 lbs | 13 | 43.33% |
| 160 – 199 lbs | 10 | 33.33% |
| 200 – 239 lbs | 1 | 3.33% |
| **English as a First Language** |  |  |
| No | 7 | 23.33% |
| Yes | 23 | 76.67% |
| **History of Mental Illness** |  |  |
| None | 25 | 83.33% |
| Anxiety, Depression, Anxiety, Suicidal Thoughts/ ideations, or History of Mental Health Illness | 5 | 16.67% |

### 3.2 Physiological indicators of Anxiety

#### 3.2.1 EDA

A one-way ANOVA based on the continuous tonic EDA data revealed that physiological arousal significantly differed across baseline, TSST, and Intervention episodes (F(2, 78) = 12.58, P <.001) as indicated in Figure 2. During baseline, the tonic EDA data ranged between 0.003 and 1.637 (M = 0.231 µS, SD = 0.355). During the TSST episode, the data ranged between 0.000 and 2.479 with the mean tonic EDA data increasing markedly (M = 0.502 µS, SD = 0.610).

**Figure 2.**
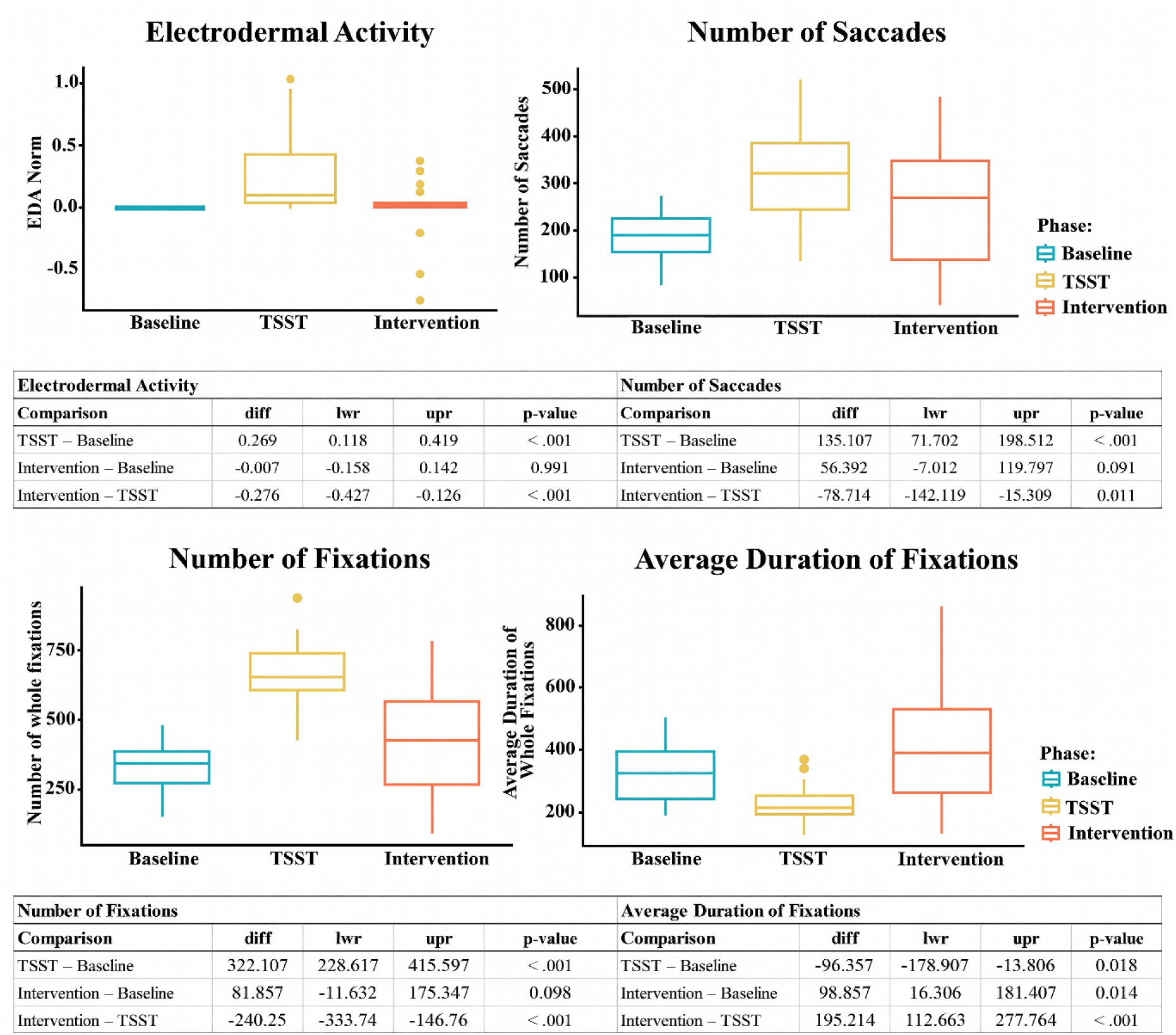
Comparisons of EDA and eye movement data across the five-minute episodes

During the intervention episode, however, the mean tonic EDA data decreased significantly (M = 0.223 µS, SD = 0.308), ranging from 0.011 and 1.417. Post-hoc pairwise comparisons indicated a statistically significant increase in mean tonic EDA levels from the baseline to TSST episodes (mean difference = 0.27, 95% CI [0.12, 0.50], p <.001) followed by a significant decrease from the TSST to intervention episodes (mean difference = - 0.28, 95% CI [-0.43,-0.13], p <.001). No statistically significant differences were observed between the baseline and intervention episodes among participants (mean difference = - 0.01, 95% CI [-0.16, 0.14], p =.991).

#### 3.2.2 Eye movements (eye fixations and saccades)

As demonstrated in Figure 2, the average fixation duration was 325.71 ms (SD = 95.33) during the baseline, 229.36 ms (SD = 57.40) during the TSST, and 424.57 ms (SD = 194.49) during the intervention episode. In addition, mean frequency of eye fixations and saccades were 334.93 (SD = 84.40) and 185.61 (SD = 51.97) during the baseline, 657.04 (SD = 115.53) and 320.71 (SD = 101.69) during the TSST episode, and 416.79 (SD = 209.59) and 242.00 (SD = 128.76) during the intervention episode. A one-way ANOVA showed a significant difference in the mean frequency of saccadic activities (F(2, 81) = 36.56, P <.001), mean frequency of eye fixations (F(2, 81) = 36.56, P <.001), and mean fixation duration (F(2, 81) = 15.94, P <.001) across the episodes. Post-hoc pairwise comparisons with adjusted p-values indicated a statistically significant increase in the mean saccade frequency from the baseline to TSST episodes (mean difference = 135.11, 95% CI [71.70, 198.51], p adj. <.001) followed by a significant reduction of mean saccade frequencies from the TSST to intervention episodes (mean difference = - 78.72, 95% CI [-142.12,-15.31], p adj. =.011). Findings revealed no statistically significant differences between the baseline and intervention episodes (mean difference = 56.39, 95% CI [-7.01, 119.80], p adj. =.091). Similarly, pairwise comparisons revealed a statistically significant increase in mean fixation frequency from the baseline to TSST episodes (mean difference = 322.11, 95% CI [228.62, 415.60], p adj. <.001) followed by a significant reduction of mean fixation frequencies from the TSST to intervention episodes (mean difference =-240.25, 95% CI [-333.74,-146.76], p adj. <.001). No statistically significant differences were observed between the baseline and intervention episodes for the average eye fixation frequencies (mean difference = 81.86, 95% CI [-11.63, 175.35], p adj. =.098). Pairwise comparisons of mean fixation durations across the episodes, on the other hand, depicted that the mean duration of fixation was significantly shorter during the TSST compared to baseline episode (mean difference =-96.38, 95% CI [-178.91,-13.81], p adj. =.018) and significantly longer in intervention episode compared to the TSST episode (mean difference = 195.21, 95% CI [112.66, 277.76], p adj. <.001). Findings also revealed a statistically significant increase in mean fixation duration from the baseline to the intervention episode (mean difference = 98.86, 95% CI [16.31, 181.41], p adj. =.014), showing a more stable visual processing in the intervention episode.

### 3.3 Subjective indicators of Anxiety

#### 3.3.1 State Anxiety Inventory Scores

Data analysis using the Friedman tests indicated that perceived anxiety levels significantly differed across baseline, TSST, and Intervention episodes (χ²(2) = 37.0, N = 25, p <.001, W = 0.74). Post-hoc Wilcoxon signed-rank tests with Bonferroni correction revealed a significant difference between baseline and TSST episodes (W = 261, p adj. =.025, r = 0.53) followed by a statistically significant decrease from the TSST to intervention episodes (W = 274, p adj. <.001, r = 0.85). Findings also revealed a statistically significant decrease in state anxiety scores from the baseline to the intervention episode (W = 325, p adj. <.001, r = 0.86).

#### 3.3.2 Semi-structured Exit Interviews

As demonstrated in Figure 3, analysis of the semi-structured interviews resulted in a total of N = 292 coded interview segments that were categorized across four major themes: Perceived Emotional Responses, Environmental Features, Telehealth Intervention Features, and Preferences. The most discussed category was Preferences with 45% of code frequency (f = 45%), followed by Environmental Features (f = 27%), Perceived Emotional Responses (f = 22%), and Telehealth Intervention Features (f = 6%).

**Figure 3.**
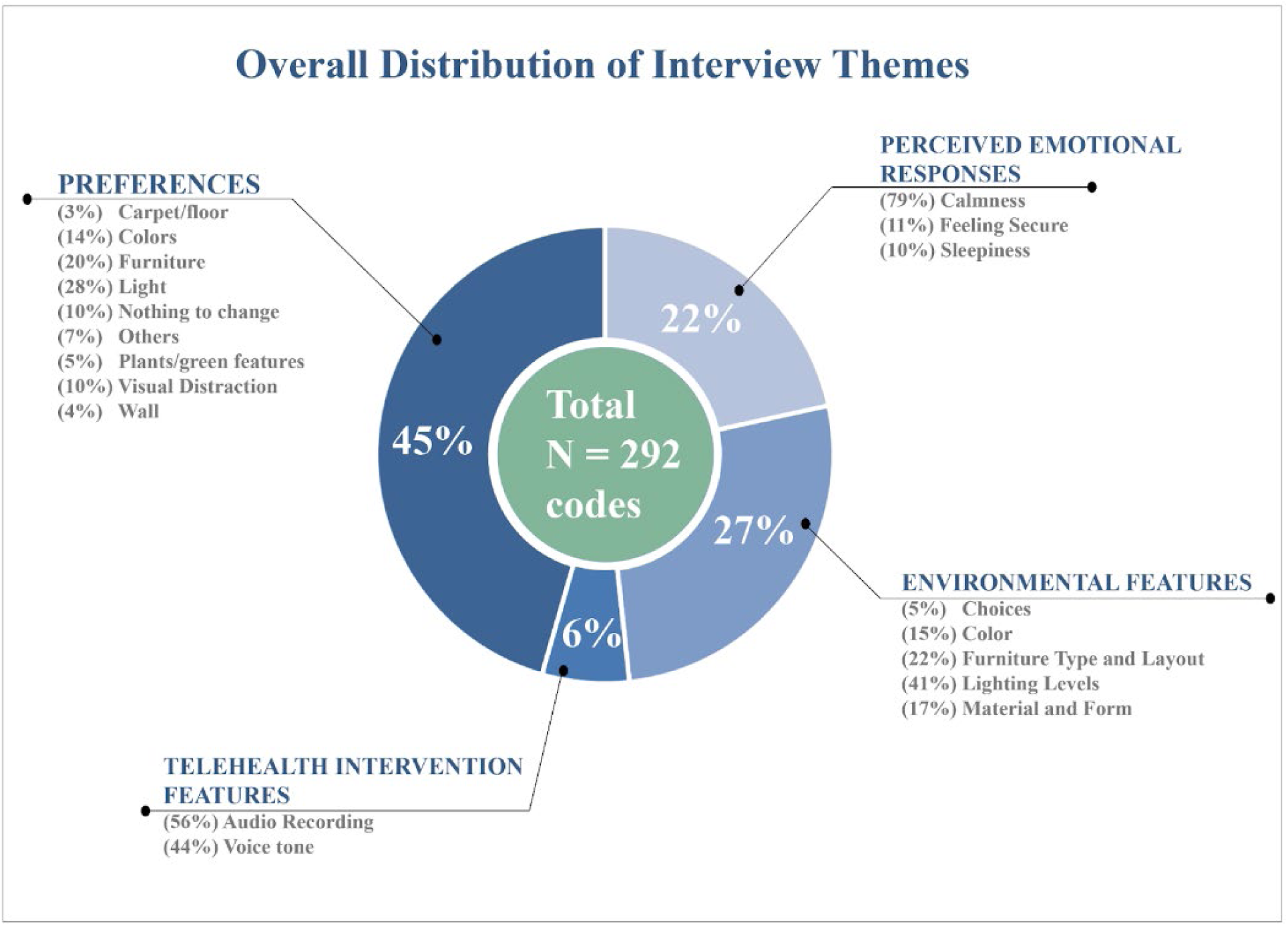
The key themes emerging from the analysis and its thematic distribution

##### 3.3.2.1 Perceived Emotional Responses

###### 3.3.2.1.1 Calmness

The participants most dominantly described their emotional responses as experiencing calmness (79%), feelings of security (11%) and sleepiness (10%). Several participants explicitly perceived space as a calming environment, making a comparison with the surrounding environment or it’s busier condition. One participant mentioned, *“It’s a very calming space, I guess. It feels very different compared to the rest of the room…just like the hustle of people moving around.”* Some participants also described various environmental features, including lighting, wall patterns, and layout, contributing to their emotional response. One participant explained,*“ I think it’s done really well, the lighting, the organization, and the wall patterns. I think they all work really well together and make it feel really relaxed”. Another* participant noted that, “*It definitely sheltered me from everything else going on, and I feel it’s a very simple room, so it lets me focus on the audio thing that was playing.”* Other participants commented on calmness in terms of telehealth-related features such as voice tone, presence of audio session, and guided meditation instructions. One participant mentioned*,“ I was originally kind of nervous during the interview part, and that meditation made me feel better after that.”* Some participants described that both the environment and the telehealth intervention played an important role in bringing a calm state. One participant stated, “*transitioning from there (the room) to the space itself and listening to the audio helped calm me down, felt relaxed, and wanted to sleep…So definitely a space you would want to go to just be in a serene nature environment, and it’s pretty much just relaxed.”*

###### 3.3.2.1.2 Safety

The participant frequently elaborated on the environment, providing a sense of safety due to privacy, enclosure, controlled access, and a sense of ownership. The participant referred to the environment their personal refuge and described it as their own kind of capsule, mentioning “*It is kind of like in a cocoon, almost I felt like I was kind of wrapped around.”* One participant highlighted that the prototype helped them feel secure, “*There’s only an entrance on one side. That helps you kind of feel more secure.”* Wall design also emerged as a contributing factor and one participant highlighted that “*the design of the walls…you don’t feel enclosed. But it’s enough where it feels private.”* Another participant added. “*I felt very calm and relaxed inside this space because the design was very private, secure.”*

#### Sleepiness

A few participants experienced sleepiness and physical relaxation within the space, noting a combination of a dark, quiet environment with telehealth audio intervention made them feel drowsy. One participant stated, “*I almost fell asleep”,* and “ *It was dark. I wanted to go to sleep.”* While another participant stated*, “I would say I felt a bit sleepy there. A bit cozy as well.”* One participant described feeling taken a nap despite spending a few minutes, *“ I feel like I just took a nap, but it was only like five minutes. So, it was cool. I liked it.”*

##### 3.3.2.2 Environmental Features

The most frequent environmental features discussed among participants included lighting quality (f = 41%), Furniture Type and Layout (f = 22%), and Materials and Form (f = 17%):

###### 3.3.2.2.1 Lighting quality

Overall, participants described lighting levels as visually appealing, composed of different light sources that shaped the sensory quality of the room. The participant described lighting quality in terms of light levels (dimness), color temperature (warm versus cool lighting), and type (soft or diffused versus harsh lights). Most participants described the lighting level as dim. One participant mentioned, “*I like the dark parts of the room. That was where my eyes went personally. Not the lighter, more engaging visuals.”* One participant also stated,*“ I really like the darkness more than anything, and it’s kind of like an enclosed space. I felt really relaxed. Just keep me focused just on the audio.” Several* participants highlighted the lighting levels were perfectly balanced. “*I think it’s really cool. It’s nice because it allows a little bit of light, and it’s not just dark. So, I think the lighting you have in there is really nice,”*. Participant also acknowledged the presence of multiple lighting sources, “*I liked all the different decorations, like the lanterns and then that stream of light (RGB LED flexible strip) on the ground.”* and another participant added that the RGB LED flexible strip provided a visual clue to focus without overwhelming the environment. *“I feel like that one (the LED flexible strip) especially gives you a little thing to focus on, but it’s not overwhelming.”* Other participants emphasized that diffused lighting features incorporated into the prototype added contrast to the space, highlighting “*I think all the little lamps around the ring and the LED strip are a nice touch. It keeps it from being too dark.”*

###### 3.3.2.2.2 Furniture Type and Layout

The participant frequently commented on variety, type, suitability, and comfort of the furniture in the multi-sensory environment. Many participants described the presence of a bean bag chair as comfortable. *“I think the green back chair is really what caught my attention, just because of how much more comforting it looked than the other ones*.” One participant further elaborated on the green chair, referencing a nice angle, posture, and curve. *“I liked the bean bag just because it’s relaxing, nice angle, but then it’s like a curve…you don’t kind of melt into it…you’re still sitting upright.”* Other added, *“I like how the furniture really is placed, and it’s not unintentional*.” And *“The furniture that was chosen to decorate and inhabit the space is fitting well.”*

###### 3.3.2.2.3 Materials and Form

Overall, participants consistently noted that the curved and circular form of the room served as a strong visual focal point and was particularly intriguing. One participant stated, *“I like the curvature of the wall. I think that’s really nice. It’s not just like 90-degree angle or anything.”* Several participants commented on the wall structure, noting*, “I liked looking at the wall because it was like weaving in and out of the panels.”* And some described it as *“well-ventilated”.* Materials such as grey acoustic PET felt panels and studs with wood patterns were considered to add warmth or contrast to the environment.

##### 3.3.2.3 Telehealth Intervention Feature

*Audio recording quality and Voice tone (guided meditation):* The audio recording was frequently referenced among the participants as a central theme of the telehealth intervention with several participants mentioning that they were “*following the instructions*”. Participants stated, *“The audio itself was pretty relaxing.”* Another participant emphasized that the combination of quietness and reassuring comments helped with the experience, “*I feel like having audio quietness and reassuring comments helps.”* or “*I listened to the audio recording, it helped calm my nervous system.”* Some participants explicitly highlighted the voice tone as a component shaping their experience, describing it as “*very soothing*” or “*calm voice*”. One participant said, *“It helped me calm down because of the way she was speaking.”* The third participant elaborated*, “Anytime I hear similar meditating, it’s always calming, and the person’s voice was really calming.”*

##### 3.3.2.4 Preferences

During the interviews, participants proposed modifications in the multi-sensory environment, including lighting features (f = 28%), furniture (f = 20%), colors (f = 14%), while other some did not want to see any changes (f=10%). Some participants also wanted certain features to be added to the environment, such as plants or green features (f = 5%), walls (f = 4%), and carpet or flooring (f = 3%), such as textured grass floors or interactive floors. Other themes (f = 7%) included preferences for personal choices such as blankets, a vase, a swing, aroma, candles, a laptop desk or a small workstation. The top three themes highlighting participants’ preferences are discussed in detail below:

###### 3.3.2.4.1 Lighting features

Participants highlighted the need to add variety of lights (e.g., fairy lights, stargazing, or lava lamps) and provide choice and control over lighting quality, such as the color temperature or light levels. One participant mentioned, *“I think it’s a little too dark, I think there should be more lights, but I don’t know if everyone would agree with that.”* While others added, *“I think for me personally, as I said, if it were a bit dimmer, more darkness, yeah, that is like a personal choice.”* or “*Personally, if I want to relax in a sensory room, I want something dark and cool like a cave.”* Participants also mentioned they wanted to see more verity of lighting features, such as candles, twinkle star, star light, or lava lamp*: “I don’t think I’d add anything else just for the sake of being able to focus on one thing, but if you add something moving like a lava lamp.”* One participant explicitly highlighted removing some lanterns: *“I feel like a couple of the lanterns could be removed, but nothing is jarring to me.”*

###### 3.3.2.4.2 Furniture

Participants expressed their different preferences related to furniture in terms of adding more comfortable seating, seating with back support or armrest, and other furniture pieces to create a homelike environment. A few participants preferred removing the furniture without a backrest. One participant stated, “*I would only find the green bean bag chair comfortable. I think that the other ones wouldn’t be very comfortable.”* Some participants indicated preference for furniture that allowed them to lay down for sensory experience, “*Maybe a chair that you lie down or a pillow on the floor…”* Another participant added, *“If a recliner were there, it would be better*!” Some participants preferred seating with a backrest or armrest, “*I feel like maybe a chair with a back on it, in case someone wants to sit upright*”. Another participant added, *“Something with an armrest, too. I think that could be pretty relaxing. Maybe if you could have one of those chairs where you could put your feet up.”* Lastly, one participant wanted to add additional furniture to create a homelike feeling in the environment,” *just small furniture to make it feel like a homey vibe in there.*”

###### 3.3.2.4.3 Colors

Several participants suggested that adding more color variations to the environment and furniture to build visual interest. One participant mentioned, *“The bean bag chair maybe could be a different color, like a soft blue or something.”* Others stated, *“I like having one grey piece of furniture. I think another piece of furniture could be like a dark blue or dark purple.” and “ Maybe add more color…these are all kinds of grey…maybe a deeper blue, or something similar to that.”* or*“ If you have more vibrant colors, but like they’re muted.”*

## 4. Discussion

### This study investigated the effectiveness of a telehealth-enabled, portable prototype with multi-sensory elements on anxiety levels among healthy young adults on a university campus

The anxiety levels were assessed via physiological (EDA, eye-fixation frequency and durations, and frequency of saccades) and subjective measures of anxiety (STAI and semi-structured interviews) during three five-minute episodes: 1) baseline, 2) exposure to stress-inducing task (TSST), 2) exposure to intervention: a multi-sensory environment with an auditory telehealth session (guided meditation).

Both physiological and subjective indicators of anxiety demonstrated that a five-minute exposure to the study intervention significantly reduced anxiety levels among participants. Specifically, the eye movement results indicated that the mean frequency of saccades and eye fixations decreased significantly when participants transitioned from the stress-inducing task to the intervention episode. Reduced saccadic frequency and fewer fixation shifts have been associated with lower levels of anxiety in prior research, suggesting diminished hypervigilant scanning. These findings are in line with reports from the existing literature. One study in a university setting investigated anxiety among students classified as anxious (N = 17) and control (N = 13) through analyzing their eye-movement patterns during e-learning activities. Based on the findings, the anxious group exhibited shorter fixation durations and higher saccade rates compared to the control group, reflecting patterns consistent with hypervigilance, attentional instability, and increased cognitive load (Agustianto et al., 2025). In contrast, fixation duration increased significantly during the intervention episode. Longer fixation durations have been linked to reduced anxiety among young adults, as individuals with lower anxiety levels exhibit less fragmented gaze behavior and fewer rapid attentional shifts (Armstrong & Olatunji, 2012). This pattern suggests that the study intervention may have facilitated sustained visual attention, consistent with a regulatory effect on anxiety levels. This result is also consistent with prior studies showing that stable eye fixations are associated with visually calming environments that promote a more relaxed psychological state (Franěk & Petružálek, 2025; Goto et al., 2025).

Electrodermal activity (EDA) findings were also consistent with these results. Tonic skin conductance levels (SCL), which reflect overall physiological arousal, decreased significantly when participants transitioned from the stress-inducing task to the intervention episode. This indicates that participants experienced lower levels of sympathetic activation while in the telehealth-enabled multi-sensory environment and highlights that the intervention helped reduce physiological arousal and overall anxiety levels. Subjective measures of anxiety from the STAI-S further supported these physiological findings. Perceived State Anxiety Inventory scores decreased significantly following exposure to the intervention, indicating that participants’ perceived anxiety levels were also reduced in parallel with objective eye-tracking and EDA measures. This result connects with existing studies where patient and staff self-reported reduction in anxiety levels and agitation behavior after use of the sensory room in the behavior healthcare setting (Dorn et al., 2020; Haig & Wagstaf, 2024; West et al., 2017). In the present study, convergence of physiological (eye-movement and EDA) and self-reported measures strengthens the validity of the findings and suggests that even brief exposure (approximately five minutes) to the proposed intervention in this study could produce measurable effects on emotional regulation and anxiety levels. These results align with existing literature demonstrating the utility of eye movement and EDA measures as indicators of anxiety-related attentional processes among individuals (Burns et al., 2025; Shechner et al., 2017).

The present study provides novel insights, demonstrating that a portable, telehealth-enabled multi-sensory environment can function as a restorative setting. While the effectiveness of multi-sensory environments have been already investigated in higher educational settings among young adults following a thirty-minute exposure to the environment (Otsuka et al., 2025), **the present study showed that a brief five-minute exposure to the telehealth-enabled multi-sensory environment as an integrated intervention could significantly reduce anxiety levels among young adults on the university campus. Importantly, the decrease in saccadic activity, eye-fixation frequencies and physiological arousal occurred alongside subjective measures of anxiety, suggesting a coordinated response among participants when exposed to the intervention.**

In addition, analysis of semi-structured interviews following exposure to all three episodes emphasized the calming effects of the proposed intervention. Comments provided by the participants delineated their preferences toward design features associated with multi-sensory environments, such as lighting quality, furniture pieces, and materials or textures. These reports are in line with the existing literature investigating the impact of multi-sensory environments on young adults. In the study by Bruce at al. (2024), students and staff in nursing educational settings indicated that improvements in lighting and interactive technology-driven tools providing choice and control for individuals could enhance the effectiveness of multi-sensory environments. This finding has been strengthened by other studies where providing lighting control to individuals improved their sensory regulation (Björkdahl et al., 2016; Hedlund Lindberg et al., 2019; Williams et al., 2023). In addition, the effectiveness of the multi-sensory environments has been shown to improve through incorporating features, such as rocking chairs, cushions, hugging pillows, blankets, and changing colored lights (Otsuka et al., 2025). These results were consistent with reports from the semi-structured interviews in the present study. Several participants elaborated preference regarding the furniture in the present study includes soft, comfortable ergonomics and having posture supporting features, such as backrest and armrest to enhance sensory experience. This observation also supported by on study exhibited that comfortable ergonomics and supporting features helped reducing distress and promote feeling of safety among young individuals in public settings (Huang et al., 2025).

### 4.1 Limitations

This study had several limitations that should be noted. First, the small sample size in a single educational setting located in the U.S. makes generalizing the results to larger populations difficult. Future studies can replicate this study with larger sample sizes to confirm the study findings. Second, the study intervention was implemented in an open lobby space of a building on the university campus where its environmental features could not be completely controlled. Third, this study did not investigate the independent effects of features in the multi-sensory environment and auditory telehealth intervention. The objective was to assess the overall effectiveness of the integrated intervention under realistic implementation conditions, not to determine the relative contribution of each individual component. Additional studies are required in a more controlled setting to investigate the impact of specific design features of the multi-sensory environment alongside other types of telehealth interventions on anxiety reduction among this population in educational settings.

## 5. Conclusion

This study investigated the effects of a brief, five-minute exposure to an integrated intervention (multi-sensory environment with telehealth) on anxiety levels among young adults on a university campus in the U.S. Results from the physiological measures, including eye movement metrics and electrodermal activity (EDA) demonstrated significant reductions in anxiety levels through decreased saccadic and fixation frequency, increased fixation duration, and reduced tonic physiological arousal among participants following exposure to the intervention. Participants also reported lower State Anxiety Inventory scores and elaborated on their calming and relaxing experience during semi-structured interviews, highlighting the alignment of findings associated with the perceived anxiety levels and physiological measures obtained through sensors. Together, these multimodal findings emphasized the value of integrating subjective and physiological measures when evaluating environmental interventions targeting anxiety. More importantly, these effects were observed after only five minutes of exposure, suggesting that brief use of the integrated intervention in educational settings may support anxiety reduction among young adults, particularly where mental health services are limited. While the present study contributes to the growing body of literature demonstrating the measurable impact of technology-enhanced multi-sensory environments, additional research is required to examine longer exposure durations, diverse populations, and other real-world contexts to generalize findings.

## Data Availability

All data produced in the present study are available upon reasonable request to the authors

## Acknowledgement

The authors would like to thank the Texas A&M Telehealth Institute, Texas A&M College of Architecture, and Stantec for their support of this study. The authors would also like to thank Sanjeela Gandhari, Texas A&M PhD student, for assistance with the data collection process.

## Ethical Considerations

Ethical approval for this study was obtained from The Texas A&M University Institutional Review Board, STUDY2025-0982. All participants provided written consent before they participated in the study.

## “Not Peer-reviewed” disclaimer

This manuscript is a preprint and has not been peer-reviewed. It should not be used to guide clinical practice.

